# Age-Standardized Comorbidity Prevalence of Cardiometabolic Conditions and Socioeconomic Inequalities in Hong Kong: A District Comparison

**DOI:** 10.64898/2026.09.24.26363851

**Authors:** Jingjing Zhou, Zhenyuan Liu, Yuchen Cai, Qiaohui Wu, David Bishai

## Abstract

**Background:** Hypertension (HTN) and Type 2 diabetes mellitus (T2DM) are leading drivers of the global cardiometabolic disease burden and major contributors to preventable morbidity and mortality. In Hong Kong, where population ageing and lifestyle-related risk factors are increasingly prevalent, both conditions and their complications pose substantial challenges to the healthcare system. Although territory-wide prevalence figures are available, small area district-level evidence on the burden of these diseases and their associated complications remains limited. Disparities are likely due to heterogeneity in demographic composition, socioeconomic status, and primary care accessibility across Hong Kong’s eighteen administrative districts. There is also a lack of accessible, age-adjusted, district-level disease visualization in Hong Kong.

**Objectives:** This study estimates the age-standardized prevalence rates (ASPR) of HTN, T2DM, and their associated complications across the eighteen districts of Hong Kong, formally evaluate the statistical significance of inter-district disparities, and rank the districts according to disease burden to inform place-based health planning.

**Methods:** This study analyzed the prevalence of chronic diseases from 2014 to 2023 in 18 districts among an adult cohort aged ≥35 years using data collected from the Hong Kong Hospital Authority (HA) electronic health records, using the International Classification of Diseases, Ninth Revision (ICD-9) diagnostic codes, the International Classification of Primary Care (ICPC) codes, and the disease flags to identify diseases. Sample weights are used to reflect the total population. HTN, T2DM, and their major complications were identified according to standardized diagnostic criteria. ASPR were calculated for each district with the WHO World Standard Population as the standard. Inter-district disparities in disease prevalence were assessed using Cochran’s Q test, with statistical significance set at *P* <0.05. The slope index of inequality (SII) was also used to analyze the socioeconomic inequality between districts. Districts were subsequently ranked according to their ASPR for each condition to identify high-burden and low-burden areas.

**Results:** 1,756,521 patients (52.71% females) contribute 9,906,729 patient-years from 2014 to 2023. Substantial inter-district variation and increasing trend were observed in the ASPR of HTN, T2DM, and their complications. The weighted ASPR of HTN varied by 27.3% from 46,701 (95% CI: 45,691-47,711) per 100,000 population in Tsuen Wan district to 59,445 (95% CI: 58,759-60,130) per 100,000 population in Yuen Long district, and the weighted ASPR of T2DM ranged from 20,464 (Tsuen Wan, 95% CI: 19,803-21,124) per 100,000 population to 28,275 (Yuen Long, 95% CI: 27,792-28,758) per 100,000 population across the districts in 2023. All diseases showed district disparities (all *P* <0.05). Cochran’s Q test demonstrated statistically significant disparities across districts and showed no convergence through the period. The SII test also showed the most serious inequality exists in HTN, with the lower socioeconomic position districts have more burden.

**Conclusion:** Significant geographic disparities exist in the prevalence of HTN, T2DM, and their complications across Hong Kong’s eighteen districts. These findings highlight the need for district-specific public health strategies, equitable allocation of primary care resources, and targeted chronic disease prevention and management programs in high-burden areas.

## Introduction

Hypertension (HTN) and Type 2 diabetes mellitus (T2DM) represent two of the most pressing chronic disease challenges confronting public health systems worldwide. These conditions form the cornerstone of the global cardiometabolic disease burden and serve as principal risk factors for cardiovascular disease (CVD), which remains the leading cause of mortality globally and contributes substantially to premature death, disability, and escalating healthcare expenditure ^1–3^.

In China, the age-standardized prevalence of HTN has risen markedly over recent decades, reaching an estimated 26.6% of adults aged 20 years and older, with a particularly striking upward trend among younger and rural populations ^4^. Evidence from neighboring Shenzhen showed variation in HTN awareness, treatment, and control across population subgroups and districts ^5–7^. In Hong Kong, where the population is rapidly ageing and lifestyle-related risk factors such as physical inactivity, dietary excess, and obesity have become increasingly prevalent, HTN and T2DM have emerged as leading drivers of morbidity. Both diseases accelerate the development of ischemic heart disease, stroke, chronic kidney disease, diabetic retinopathy, and lower-limb vascular disease. Focusing on this patient population offers a meaningful lens to evaluate disparities in chronic disease management in Hong Kong.

Territory-wide prevalence estimates provide a useful overview ^8^, but mask small area disparities. Geographic disparities in cardiometabolic disease burden and healthcare utilization are shaped not only by individual-level characteristics but also by structural attributes of place, including the density and capacity of primary care services, socioeconomic development, urbanization, and demographic composition ^1,9,10^. Studies in mainland China and Taiwan have shown that even within a single jurisdiction operating under universal health coverage, marked inter-district differences in disease prevalence, hospitalization, and outcomes persist, often reflecting underlying inequities in primary care access, socioeconomic status, and the urban–rural divide ^9,11,12^. Hong Kong’s eighteen administrative districts likewise differ in their demographic composition, socioeconomic profile, and built environment. As an exclusively urban area, the density of healthcare facilities, and accessibility to primary care services has only muted variability. Social heterogeneity can translate into meaningful disparities in disease burden, even within a geographically compact city. District-level comparison enables the identification of high-risk localities where targeted public health interventions and resource reallocation may yield the greatest impact, as has been demonstrated by district- and community-level analyses of HTN management in Shenzhen ^7,8^. Second, this approach allows the recognition of positive deviants that can reveal resilience factors ^9^. Third, examining geographic disparities in HTN, T2DM, and their complications can illuminate the social and structural determinants of health that operate beyond the individual clinical encounter, supporting place-based approaches to chronic disease control ^1,13^. Despite the value of such analyses, comprehensive district-level evidence on the prevalence of HTN, T2DM, and their associated complications in Hong Kong remains limited.

The purpose of this study is therefore to estimate the age-standardized prevalence rates (ASPR) of HTN, T2DM, and their associated complications across the eighteen districts of Hong Kong, to formally assess the statistical significance of inter-district disparities, and to rank the districts according to disease burden. By doing so, we aim to inform district-specific health planning, guide the equitable allocation of primary care resources, and support evidence-based strategies for place-based chronic disease prevention and control in Hong Kong.

## Methods

### Study Design and Population

We analyzed the prevalence of chronic comorbidities among adults aged ≥35 years with T2DM and/or HTN, using electronic health records from the Hong Kong Hospital Authority (HA). Because a case can only be identified in someone who attends HA services, and attendance varies by age, HA records under-ascertain true population prevalence. For the general-population analysis we corrected for this with a district-level inverse-attendance weight, applied before estimating prevalence and age-standardized rates^14^.

### Disease Identification from Multiple Data Sources

Disease status was drawn from four administrative databases, each using a different clinical coding system. Inpatient (IP) and outpatient (OP) records identified 13 chronic diseases via the International Classification of Diseases, Ninth Revision (ICD-9) codes; the primary care database identified 11 diseases via ICPC codes; and the Disease Management and Chronic Services (DMCS) database identified a further 8 diseases via status flags rather than diagnostic codes ^15,16^. Supplementary Table 1 lists the codes and flag definitions used for each disease across the data sources.

### Record Linkage and Harmonization

Linkage proceeded in stages. First, the IP, OP, and ICPC datasets were merged at the patient level. For patients appearing in more than one, each disease’s onset date was set to the earliest recorded date across sources, so onset reflects the first documented encounter regardless of care setting.

The combined dataset was then merged with DMCS to form a unified analytic file spanning all four sources. Because the conditions studied are chronic and irreversible, we applied a carry-forward rule: once a patient was recorded with a disease in any source in any year, they were classified as having it in all later years. This prevents misclassifying patients as disease-free in years with no healthcare encounter.

### Demographic and Geographic Linkage

Sex and year of birth were linked to the analytic file, with year of birth used to derive age for each calendar year.

Residential district was derived from three encounter databases: General Outpatient (GOP) attendance, IP admission, and Specialist Outpatient (SOP) attendance. Records were pooled, and for each patient-year the most frequently recorded district was assigned, producing a patient–district–year mapping that was then linked to the analytic file.

For patients still missing a district, we imputed within each patient by carrying forward or backward the nearest known district within a three-year window, assuming the residential district is stable over short periods. When both a prior and a subsequent known district fell within this window, the temporally closer one was used. Patients with no district recorded within three years remained unassigned and were dropped.

### Deduplication and Aggregation

The linked file was deduplicated to one record per patient per calendar year. From this patient-year file we tabulated, for each combination of year, district, age group, and sex, the number of patients with each disease. Throughout, strata are indexed by age group *a*, district *d* (*κ* = 18 districts in Hong Kong), calendar year *t*, and sex *s*.

We computed parallel set of ASPR using data frame of aggregated counts by year, district, sex (both/male/female), and age group (35-44, 45-54, 55-64, 65+), each carrying its stratum-specific denominator in census population and case count from HA records.

### Estimation of Age-Standardized Prevalence

To compare prevalence over time while controlling for age structure, we age-standardized all rates using WHO standard weights *w_a_* ^17^. The five-year WHO weights were aggregated to our four age groups and rescaled to sum to one; A single fixed standard was used across all years so temporal trends are not confounded by a changing standard. Two distinct estimation approaches were used so that every stratum retained adequate case numbers (cases > 5) ^18^. We used a direct standardization (approach 1) for high-prevalence conditions (like T2DM, HTN, lipid disorder, and retinopathy) ^19^. We also used a Bayesian hierarchical model (approach 2) for low-prevalence conditions (heart failure, stroke, atrial fibrillation, cataract, coronary heart disease, peripheral vascular disease, renal failure, skin ulcer or amputation of lower limb, and neuropathy). This model was fitted via integrated nested Laplace approximation, INLA ^20,21^ to stabilize estimates and yield full posterior credible intervals.

Because cases arise only among HA attendees, a crude rate against the resident population under-counts true prevalence. For each stratum we therefore defined an inverse-attendance weight

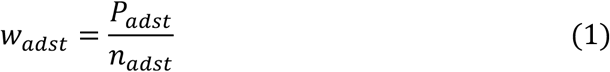

where *P_adst_* is the census resident population and *n_adst_* is the number of HA attendees, so each attendee represents *P*/*n* residents. This weight was applied only when the denominator was the resident population.

### Approach 1: direct standardization

Let *c_adst_* be the comorbid case count and 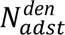 be the district residents over 35. The crude age-specific prevalence is

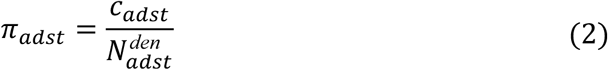

Applying the sampling weight scales observed cases to the resident population, giving the weighted age-specific rate

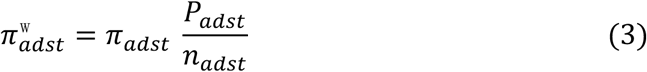

The ASPR (per 100 000) ^22^ is the age-weighted sum

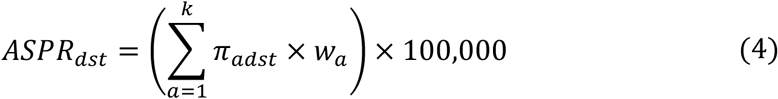

### Approach 2: Bayesian hierarchical model

To stabilize small strata and propagate uncertainty into the standardized rates, we fitted a Bayesian model using INLA, separately for each year and sex. Stratum case counts were modelled as binomial with a logit link,

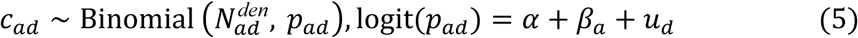

Here 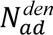 denotes the population denominator within age group *a* and district *d*. The parameter *p_ad_* represents the underlying true prevalence proportion specific to that stratum. To model this probability, α is the intercept, *β_a_* are fixed age-group effects, and *u_d_* is a district random effect, *u_d_* ∼ N(0, *σ*^2^), which lets each district borrow strength from the others. We placed a penalized-complexity prior on the district standard deviation *σ*, with Pr(*σ* > 1) = 0.01, and zero-case strata were retained in the analysis to avoid introducing bias.

From each model we drew 1 000 posterior samples. For each sample, we converted the posterior prevalences *p_ad_* to rates per 100 000 and standardized with the WHO weights to yield posterior distributions, summarized as posterior medians with 95% credible intervals (2.5th and 97.5th percentiles).

For the general-population analysis, each crude rate was multiplied by the inverse-attendance multiplier *P*/*n* before standardization, propagating uncertainty through the same draws ^20,21^.

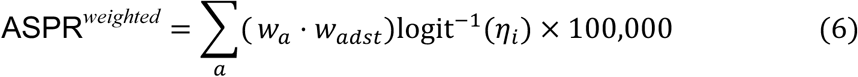

which reduces to ∑*_a_ w_a_* (cases*_a_*/*n_a_*) and therefore targets the same quantity as the direct-method weighted ASPR, while additionally yielding full posterior credible intervals.

Standard errors were estimated treating age-specific counts as independent Poisson variables,

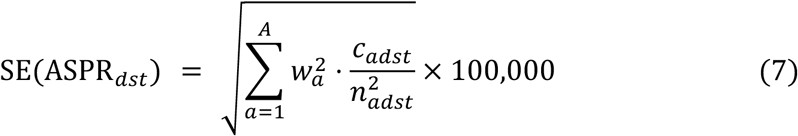

with 95% confidence intervals as *ASPR* ± 1.96 ⋅ *SE*. Where age-stratum case counts were small, gamma-distribution-based intervals yielded equivalent results.

### Quantifying between-district inequality

For each disease-year-sex stratum we summarized the distribution of ASPR across the *k* districts using descriptive statistics (mean, standard deviation, median, interquartile range, range) and two unweighted inequality measures. The Gini coefficient ^23^ was computed as

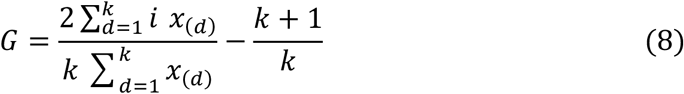

where *x*_(*d*)_ denotes the *d*-th order statistic of the district ASPR; *G* = 0 indicates perfect equality and larger values indicate greater inequality. The Index of Disparity was computed as the mean absolute deviation from the cross-district mean, expressed as a percentage of that mean:

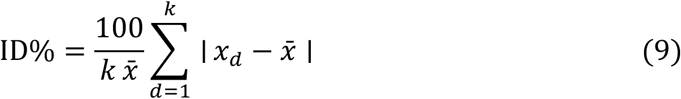

where the deviations are taken about the cross-district mean 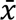 and normalized by *k*; this mean-referenced variant was adopted in preference to the reference-rate formulation to remain consistent with the unweighted descriptive statistics reported throughout.

### Test of between-district heterogeneity

To test whether between-district variation in ASPR exceeded sampling variability, we applied Cochran’s Q test ^24^ in a meta-analytic framework, treating each district’s ASPR as a precision-weighted estimate. Let *r_d_* and *se_d_* be the ASPR and its standard error in district *d*(*d* = 1, …, *k*), and let 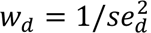. The inverse-variance-weighted overall mean is

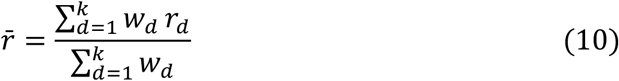

and

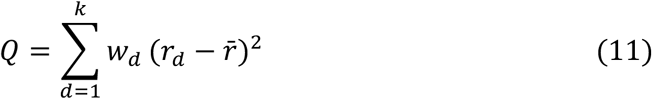

which under the null hypothesis of homogeneity asymptotically follows a *χ*^2^ distribution with *k* − 1 degrees of freedom. We report the Q statistic, degrees of freedom, and the upper-tail *P* value, taking *P* < 0.05 evidence of heterogeneity.

To quantify the *magnitude* of heterogeneity, we report

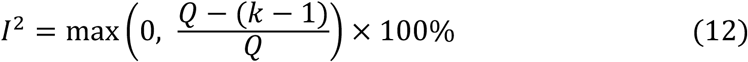

the percentage of total variation in district ASPR attributable to true between-district heterogeneity rather than sampling error. Following conventional, values around 25%, 50%, and 75% were interpreted as low, moderate, and high heterogeneity, respectively.

The Cochran’s Q framework was preferred over a chi-square test of independence because it explicitly incorporates the precision of each district’s estimate through inverse-variance weighting, thereby down-weighting estimates from small districts with wide confidence intervals. It also operates directly on age-standardized rates and is therefore consistent with the descriptive quantities reported throughout the paper.

### Socioeconomical inequalities

District-level socioeconomic indicators and mid-year resident population were obtained from the Hong Kong Census and Statistics Department District Council district data tables ^25^. The district is the smallest administrative unit for which the required socioeconomic and health indicators are consistently and publicly available. Our primary measure of area-level socioeconomic position was a Socioeconomic Index (SEI) derived by principal component analysis (PCA), following the general approach of established neighborhood socioeconomic and deprivation indices. Candidate indicators were screened for multicollinearity, and five non-redundant variables were retained: average household size; median monthly household income of economically active households, excluding foreign domestic helpers; the percentage of the population aged 15 and over with post-secondary education; the percentage of the population who were full-time students; and the percentage of the population who were homemakers.

Each variable was standardized to zero mean and unit variance across all 180 district-years, and PCA was performed on the resulting correlation matrix. The first principal component (PC1) was retained as the SEI and oriented so that median household income loaded positively so that higher scores denoted higher socioeconomic position. We did not impose a priori signs on the input variables; the direction of each variable’s contribution was determined by the data. PC1 scores were rescaled to a 0–100 range across the Hong Kong district-years.

Absolute socioeconomic inequality in ASPR was quantified using the Slope Index of Inequality (SII), estimated by the regression-based method of Pamuk ^26^. Districts were ranked from lowest to highest socioeconomic position according to the index under consideration, and each district was assigned a fractional rank equal to the midpoint of its cumulative share of the total population when districts are ordered by socioeconomic position (values in the open interval 0–1). ASPR was regressed on the fractional rank by weighted least squares, with weights equal to district total population. The fitted slope is the SII and the modelled absolute difference in ASPR (per 100,000) between the highest- and lowest-ranked extremes of the socioeconomic scale. A negative SII indicates higher prevalence in lower socioeconomic-position districts.

### Subgroup and sensitivity analyses

All analyses were repeated for males, females, and both sexes combined, and stratified by year to examine temporal stability of the between-district heterogeneity. Two outlier district-disease combinations (neuropathy and renal failure in Sham Shui Po) with known data-quality issues were masked from heterogeneity calculations. To assess the robustness of the findings to the operationalization of socioeconomic position, we repeated all analyses using three alternative rankings: median household income alone, representing the material dimension; post-secondary education alone, representing the educational dimension; and a two-component Socio-Demographic Index (SDI-2), defined as the geometric mean of income and education after each was rescaled to the interval [0.01, 1] using global minima and maxima. Income and education thus served both as stand-alone rankings (the two constituent single-variable gradients) and as inputs to the composite indices.

Data cleaning was performed using Stata 18.5 ^27^ on the university secure platform because of data privacy requirements. ASPR and disparities analyses used R 4.4.1 ^28^ with tidyverse ^29^, ggplot2 ^30^, and DT ^31^ packages; the interactive dashboard was built in Shiny ^32^ and plotly ^33^. Robust regression lines shown in figures were fitted by iterated re-weighted least squares (MASS::rlm) ^34^; reported SII estimates are from population-weighted least squares. Two-sided *P* values < 0.05 were considered statistically significant.

## Results

### Study Population

The analytic file comprised 1,756,521 unique patients aged ≥35 years contributing 9,906,729 patient-years across the study period (2014 - 2023) in Hong Kong. The cohort was 52.71% female, and the age distribution shifted toward older groups over the study period, with the proportion aged 65+ years rising from 53.73% to 63.42%. Following the two-step sequential imputation, district could be assigned for 99.44% of patient-years; 0.56% remained unassigned and were excluded from the district-level analyses.

### Age-standardized prevalence in the general population

The weighted ASPR of cell diseases increased across the study period. The highest prevalence condition was HTN, with the weighted ASPR increased from 40,517 per 100,000 (95% Confidence Interval [CI: 40,385-40650]) in 2014 to 54,026 per 100,000 (95% CI: 53,821-54,230) in 2023, an absolute increase of 13,508 per 100,000 (a relative increase of 33.3%), followed by lipid disorder (from 25,108 to 40,964 per 100,000; a relative increase of 63.2%) and T2DM (from 17,372 to 24,569 per 100,000; a relative increase of 41.4%). Shown in Table 1, Figure 1-2, and Supplementary Dashboard.

**Table 1.**
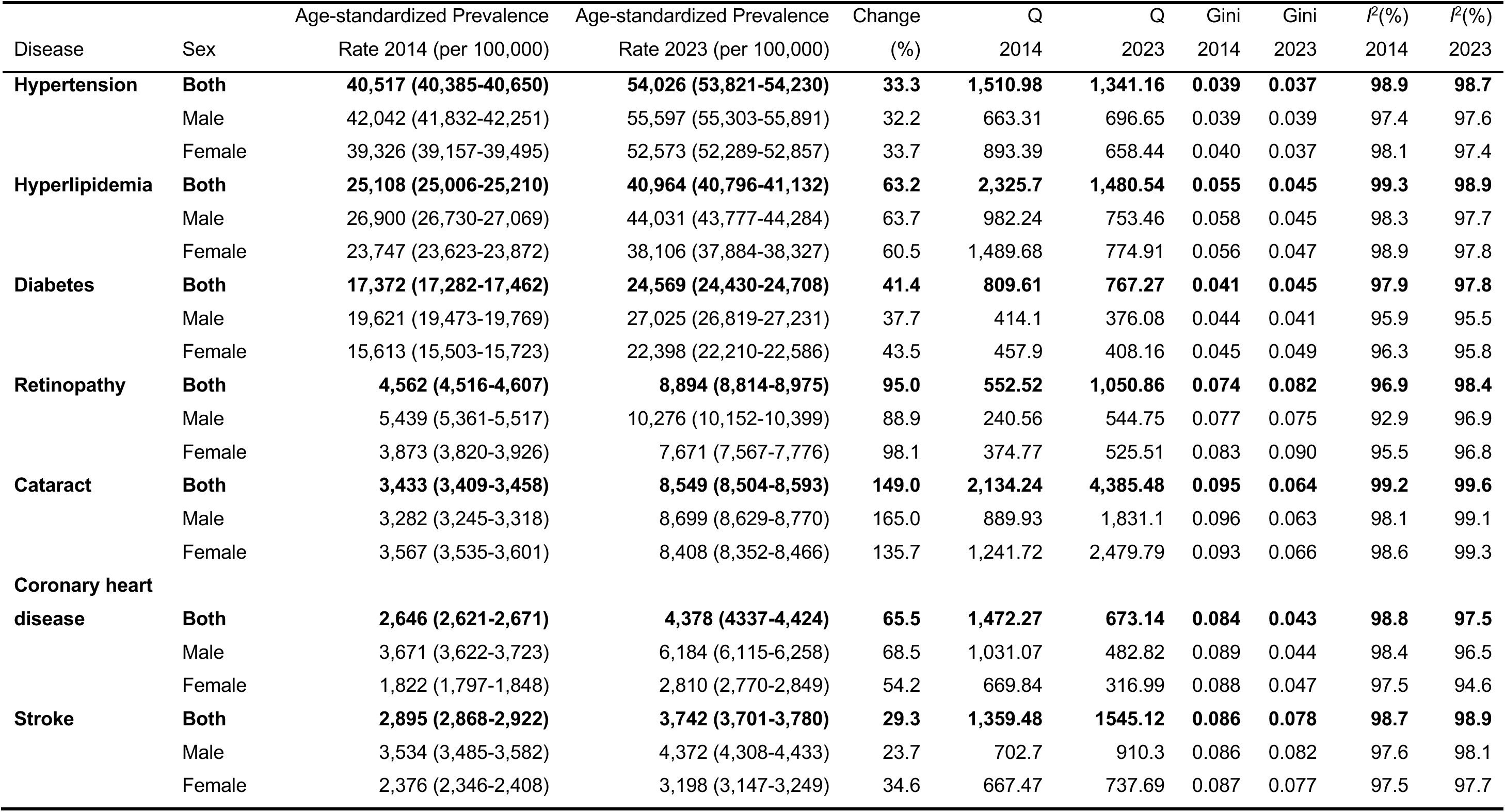
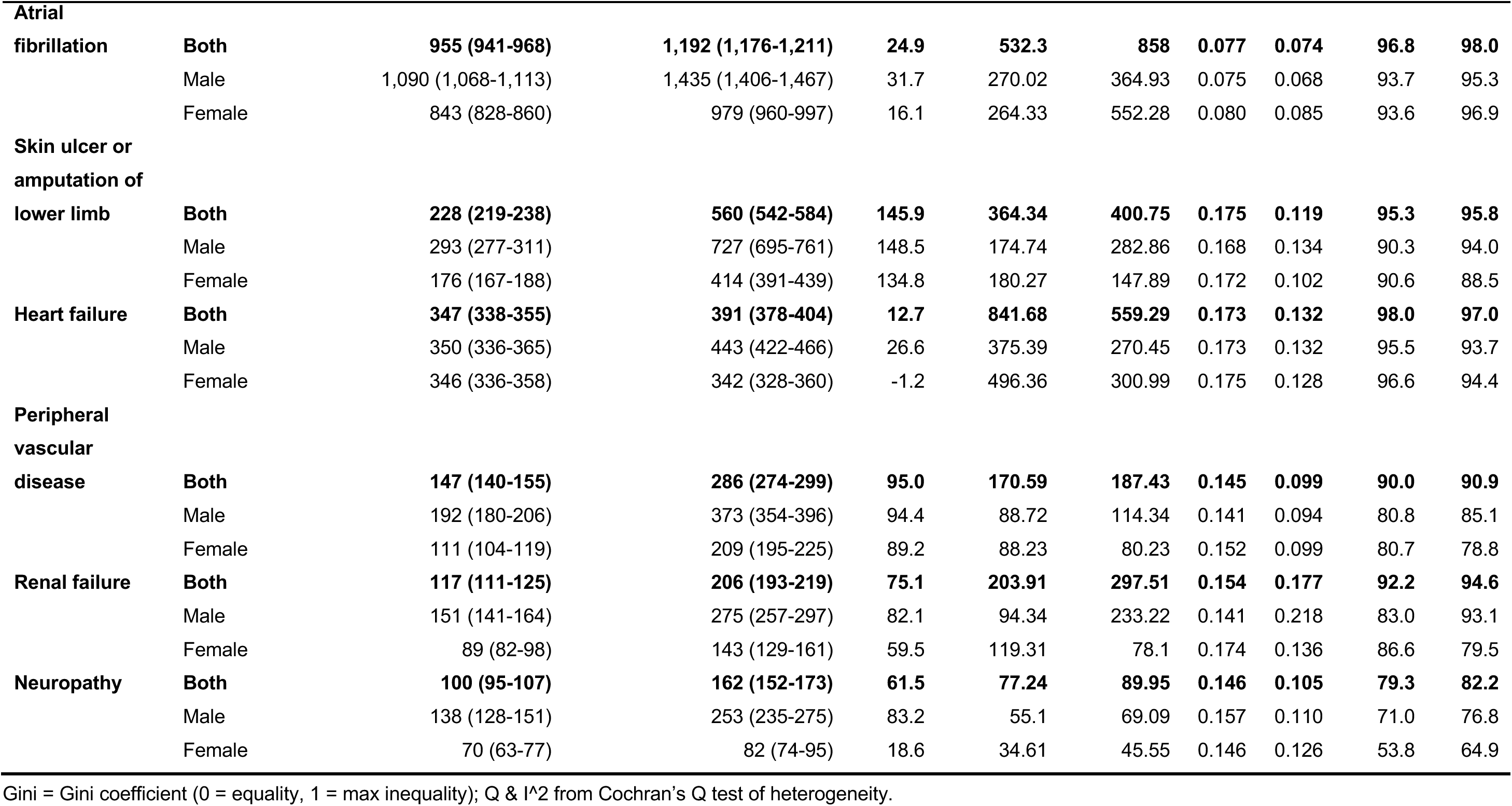
Age-standardized Prevalence Rate of Diseases in Total Population.

**Figure 1.**
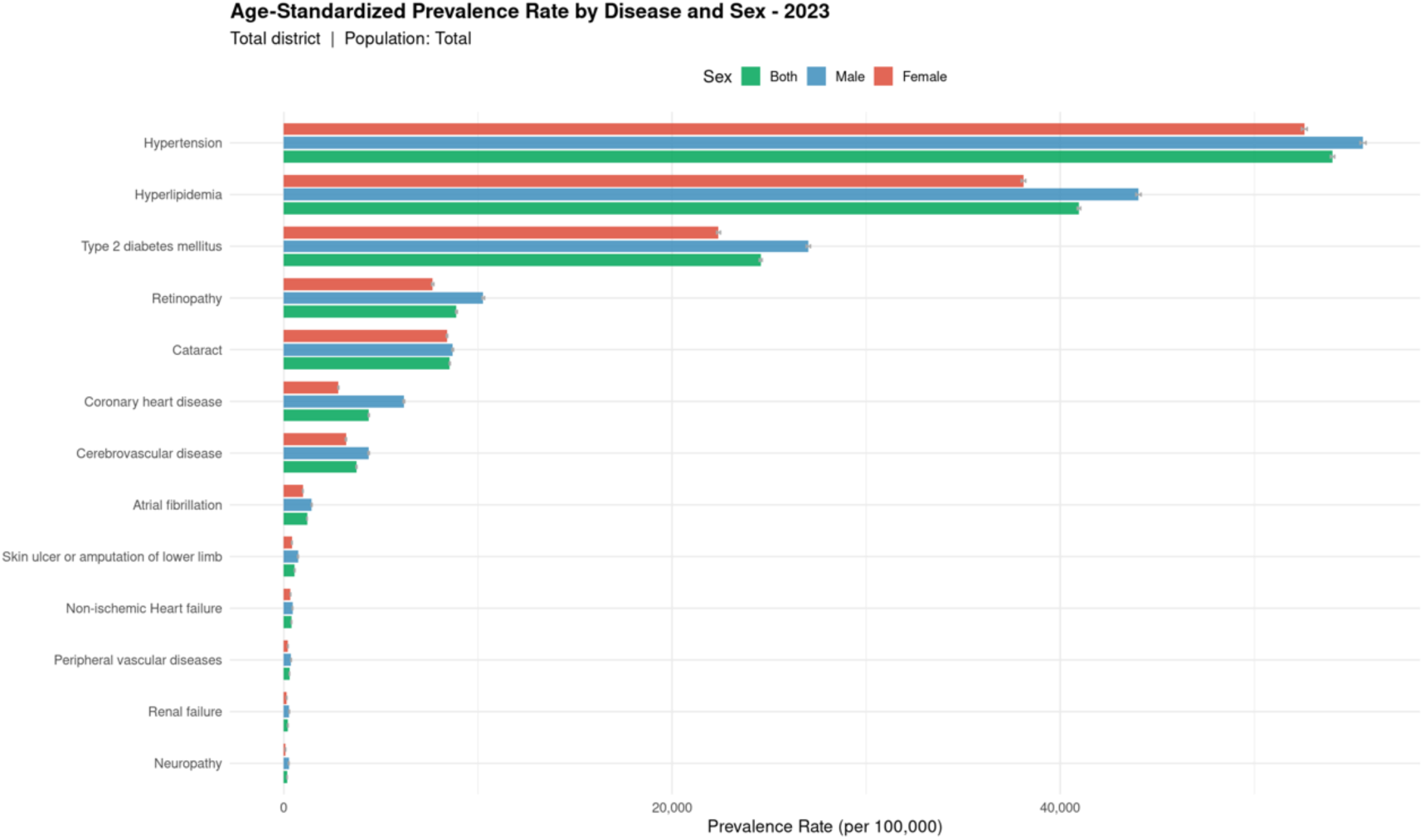
The Weighted Age-standardized Prevalence Rate of Diseases in Total Population.

**Figure 2.**
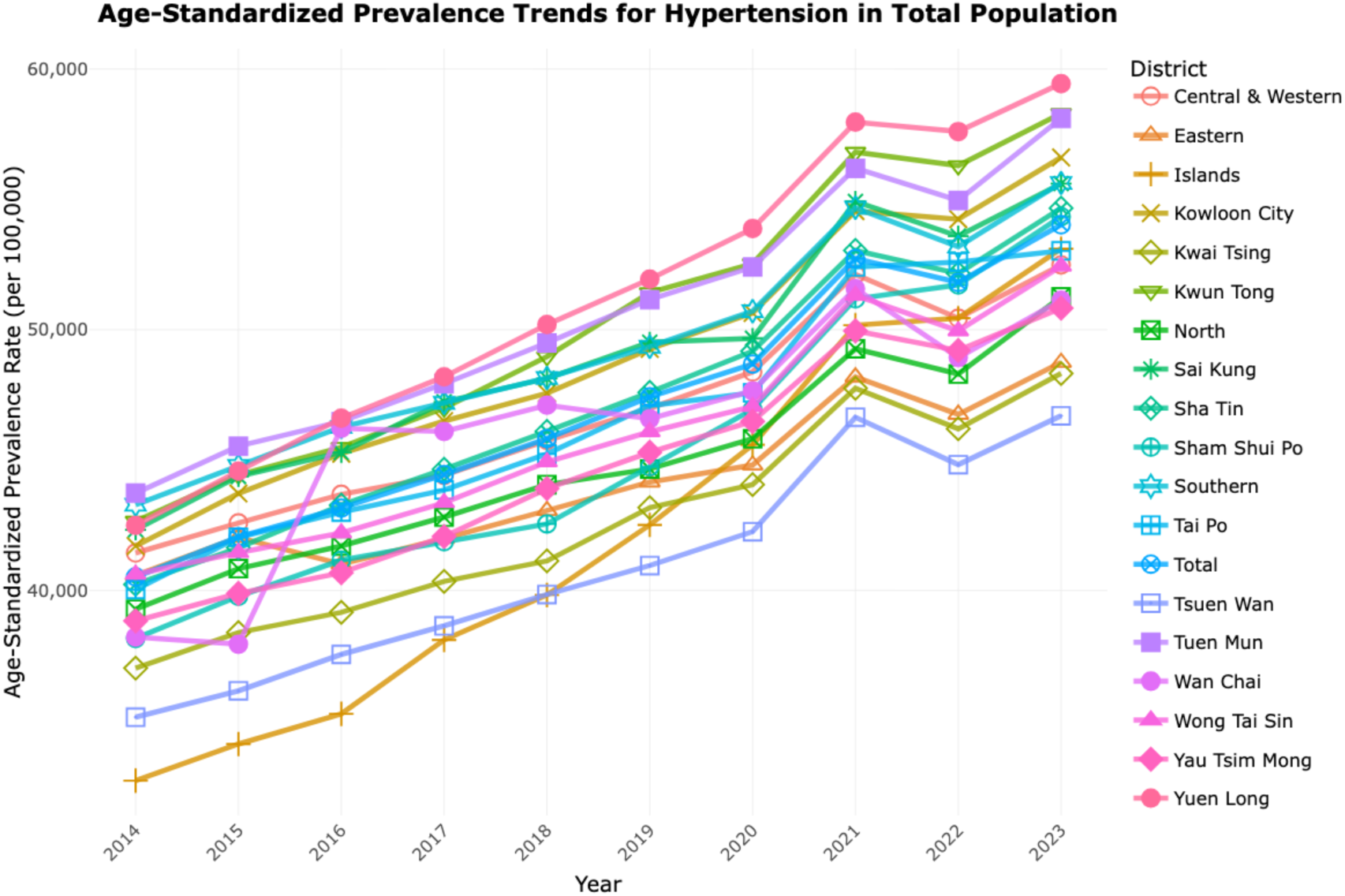
The Weighted Age-standardized Prevalence Rate of Hypertension in Total Population.

### Between-district inequality

The magnitude of geographic inequality varied substantially by condition but followed a consistent structure. HTN and CHD were the most evenly distributed (spatial Gini 0.037–0.041), while renal failure was the most geographically concentrated (Gini 0.177–0.190). The Index of Disparity reproduced this same ordering and is reported in full in Tables 1 and online Dashboard. To illustrate the spread, in 2023 the weighted ASPR of HTN in the total population ranged 27.3% across districts, from 46,701 (95% CI: 45,691-47,711) per 100,000 in Tsuen Wan to 59,445 (95% CI: 58,759-60,130) per 100,000 in Yuen Long, Shown in Figure 2. The weighted ASPR of T2DM ranged from 20,464 (Tsuen Wan, 95% CI: 19,803-21,124) to 28,275 (Yuen Long, 95% CI: 27,792-28,758) per 100,000 across the eighteen districts.

District rankings were stable over time. Yuen Long, Southern, and Wong Tai Sin persisted among the highest-burden districts and Tsuen Wan and Tuen Mun among the lowest, with no single district dominating the high-burden profile across conditions.

### Failure to achieve district convergence

The central finding is that disparities show few signs of converging. Between-district heterogeneity was statistically significant for every condition in 2023 (Q = 89.95-4,385.48, all *P* < 0.001). More strikingly, every condition remained in the high-heterogeneity category throughout the decade. *I*^2^ ranged from 82.2% (neuropathy) to 99.6% (cataract) in 2023, and no complication transitioned out of high heterogeneity between 2014 and 2023. See Table 1.

This persistence held even where the crude gaps were narrowing. Absolute inequality metrics did contract. The inter-district range in coronary heart disease ASPR halved from 67.0% in 2014 to 35.5% in 2023, with the Gini falling from 0.084 to 0.043, yet *I*^2^ for the same conditions did not fall and in several cases rose. Cataract, the most heterogeneous complication, illustrates the pattern most sharply. In other words, as prevalence and precision rose, even the shrinking absolute differences between districts remained statistically large, convergence in burden was not matched by convergence in the underlying geographic structure. A decade on, and after correction for age structure, Hong Kong’s districts remain divergent. Heterogeneity estimates were unchanged when the two masked outlier strata were omitted. See Table 1.

### The socioeconomic index

The first principal component of the five multicollinearity-screened indicators accounted for 59.7% of the total variance and provided a coherent one-dimensional summary of area-level socioeconomic position. Oriented to income, the data-driven loadings were substantively interpretable: median household income and post-secondary education loaded positively, while household size, the proportion of homemakers and the proportion of full-time students loaded negatively.

### Socioeconomic inequalities in age-standardized prevalence rate

Ranking districts by the PCA-SEI, the largest gradients in 2023 were negative for the cardiometabolic conditions, indicating higher prevalence concentrated in lower socioeconomic-position districts (Table 2; Figure 3). The steepest SII gradients were observed for HTN (SII −5,620 per 100,000), T2DM (−3,955), hyperlipidemia (−3,576) and retinopathy (−2,684). In relative terms, ASPR at the bottom of the socioeconomic scale exceeded that at the top by an estimated 8–26% for these four conditions. Conditions with cardiac or vascular presentations showed shallow SII gradients in the opposite direction, with somewhat higher recorded prevalence in higher socioeconomic-position districts, though none reached significance: coronary heart disease (SII +395), atrial fibrillation (+148), heart failure (+69) and renal failure (+69). Stroke (-425), neuropathy (-53), cataract (-131), and peripheral vascular disease (-9) showed SII gradients close to zero.

**Table 2.** Slope Index of Inequality by condition, 2014 and 2023, both sexes, PCA-SEI ranking.

| Disease | Slope Index of Inequality 2014 (95% CI) | Slope Index of Inequality 2023 (95% CI) |
| --- | --- | --- |
| Atrial fibrillation | 168 (-55 to 390) | 148 (-148 to 443) |
| Cataract | 301 (-648 to 1,250) | -131 (-2,010 to 1,749) |
| Coronary heart disease | 435(-280 to 1,151) | 395 (-228 to 1,018) |
| Heart failure | 54 (-129 to 237) | 69 (-90 to 229) |
| Hypertension | -741 (-5,195 to 3,714) | -5,620 (-11766 to 526) |
| Hyperlipidemia | 953 (-3,443 to 5,348) | -3,576 (-9,202 to 2,049) |
| Neuropathy | 11 (-28 to 49) | -53 (-94 to -11) * |
| Peripheral vascular disease | -11 (-77 to 56) | -9 (-102 to 84) |
| Retinopathy | 325 (-647 to 1,297) | -2,684 (-4,697 to -670) * |
| Renal failure | 39 (-65 to 144) | 69 (-89 to 226) |
| Stroke | -452 (-1,131 to 227) | -425 (-1,261 to 412) |
| Diabetes | -565 (-2,850 to 1,720) | -3,955 (-6,882 to -1,028) * |
| Skin ulcer or amputation of lower limb | 55 (-106 to 217) | -34 (-257 to 188) |

\* $P < 0.05$ ; CI: Confidence interval

**Figure 3.**
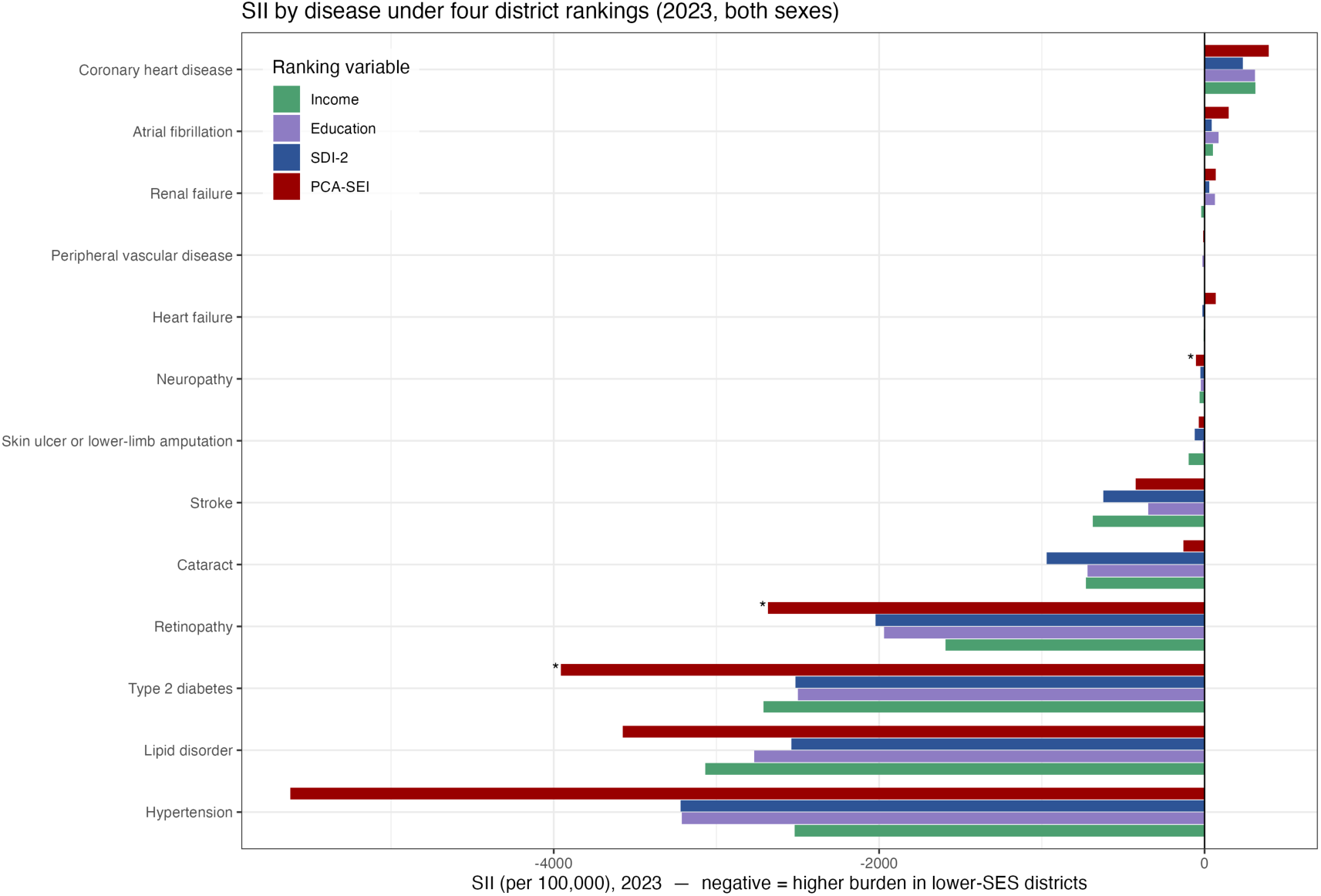
Slope index of inequality by condition under four district rankings for both sexes in 2023 * *P* < 0.05.

Where gradients were present, they generally steepened over the study period. For the four conditions with the largest 2023 gradients, the SII moved from near zero or positive in 2014 to strongly negative by 2023: HTN from −741 to −5,620 (Figure 4), T2DM from −565 to −3, 955, hyperlipidemia from +953 to −3,576, and retinopathy from +325 to −2,684. This indicates a widening of absolute socioeconomic inequality in the recorded prevalence of these conditions over the decade.

**Figure 4.**
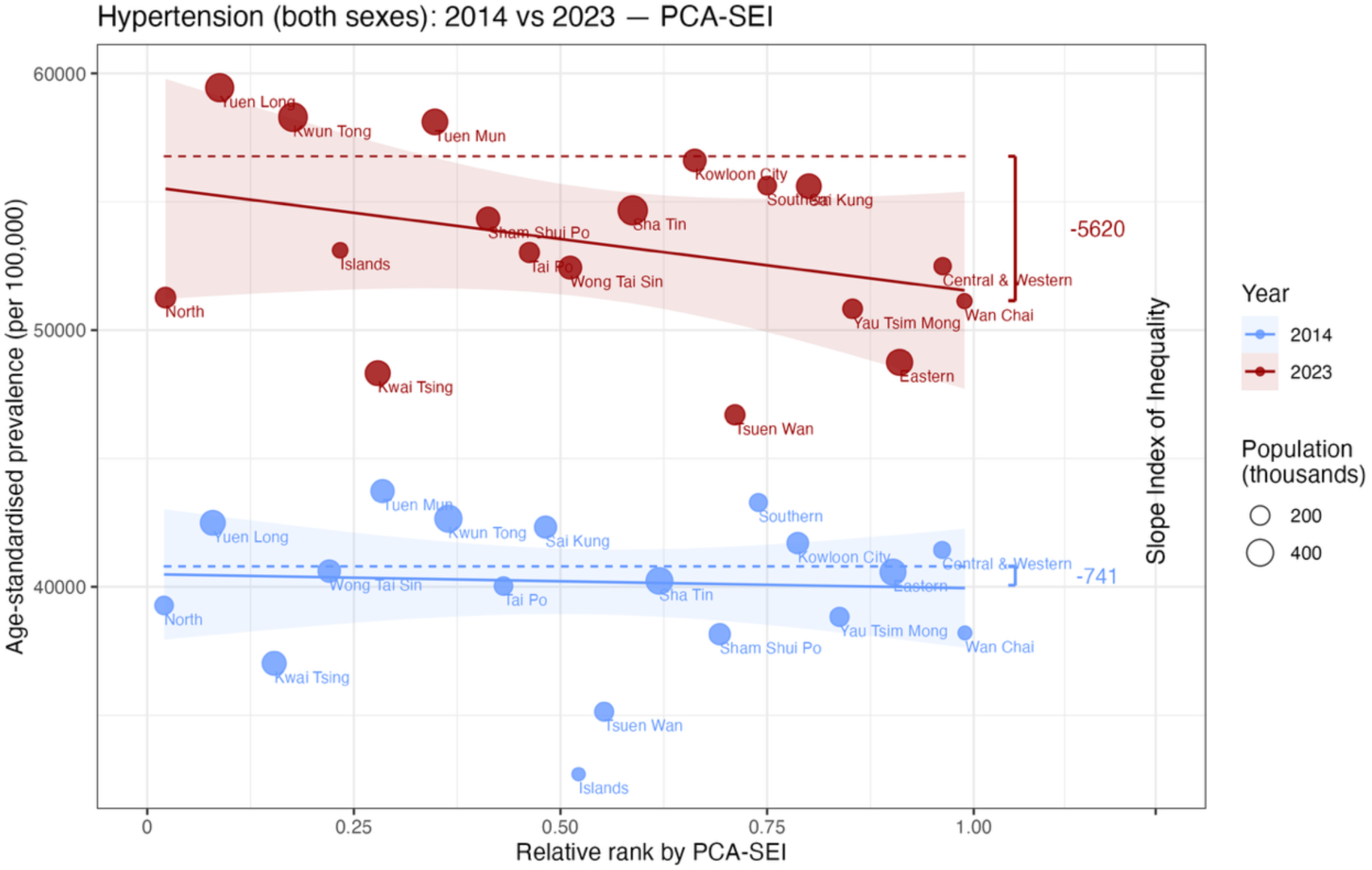
Slope index of inequality by hypertension for both sexes in 2023 95% Confidence interval: -5,195 to +3,714 (2014); -11,766 to +526 (2023)

Ranking districts by income alone and by education alone produced gradients in the same direction as the composite index for every condition with a material gradient. Both dimensions identified the same conditions as socioeconomically patterned, indicating that the inequalities are driven by socioeconomic position broadly rather than by any single indicator.

SII estimates were highly concordant across the four rankings. The Pearson correlation between SII estimates (both sexes, all strata) was 0.925 for SDI-2 versus the PCA-SEI, 0.912 for education versus PCA-SEI, and 0.856 for income versus the PCA-SEI. Where a gradient existed, all four measures detected it in the same direction and of similar magnitude; disagreements in sign occurred only for conditions whose SII was close to zero, where small differences in district ordering can reverse a negligible slope. The socioeconomic gradients we report are therefore not an artefact of a particular index.

## Discussions

This study quantified the prevalence and the geographic distribution of HTN, T2DM, and their complications among Hong Kong adults aged ≥35 years between 2014 and 2023. The ASPR of HTN, T2DM, and their complications rose steadily over the decade, showing a rising burden of cardiometabolic disease independent of population aging ^2,35^. Substantial inter-district variation was evident, with the magnitude of inequality differing markedly by condition. The low-prevalence conditions (renal failure, heart failure, neuropathy, skin ulcer or lower-limb amputation) consistently occupied the high spatial inequality end of the distribution, whereas common conditions (HTN, T2DM, CHD, retinopathy, stroke) were among the most evenly distributed. This ordering held across the population.

The persistence of significant between-district heterogeneity after age standardization, as shown by Q, indicates that the observed inequality cannot be attributed to differences in district age structure. The exceptionally large Q value for cataract in all three analyses reflects both the high case counts available for this common ophthalmic condition, which confer high precision and therefore high power to detect even modest absolute differences, and genuine variation in its district-level burden.

Our results show a failure to converge over time. Even where the absolute gaps in some cases narrowed over the decade, the relative statistical heterogeneity quantified by *I*^2^ remained high throughout, with no complication transitioning out of the high category and the least-dispersed conditions still exceeding 82%. This is not the reassuring picture of a system closing its geographic gaps. It is a decade in which the shape of inequality proved stubbornly fixed even as the disease burden itself grew. As prevalence and case counts rose, our estimates became more precise, so even the smaller absolute differences that remained were still large relative to their sampling error. Narrowing ranges therefore coexisted with undiminished and sometimes rising. We should take from this that after a decade, and after full adjustment for age, its districts remain as statistically distinct as ever. Our study aligns with the theoretical framework of Victora et al. ^36^, who posits that inequity gaps begin to close only after affluent districts have reached a ceiling of health improvement. Our findings suggest that this saturation point has likely not yet been achieved in the studied districts; consequently, the expected narrowing of the disease burden gap remains pending, emphasizing that convergence is an inherently delayed and long-term outcome. Moreover, while the inverse equity hypothesis predicts eventual convergence, this process is perpetually disrupted by the continuous introduction of new health technologies. The persistent divergence may reflect a ‘moving target’ phenomenon: wealthier districts consistently adopt newer, more effective interventions (e.g., advanced diagnostics or treatments) before poorer ones, thereby offsetting the catch-up progress of disadvantaged areas and preventing the disease burden gap from narrowing.

A single high *I*^2^ could theoretically coexist with a shift in the rank ordering, but the same districts occupied the extremes year after year. Heterogeneity directed at the same districts over ten years points to a stable, structural feature of Hong Kong’s disease geography. Plausible drivers, including socioeconomic composition ^37^, local service availability ^38^, cultural influence or patients behavior ^39^, cross-city hospital utilization ^40^, coding norms for health services ^41^, or biological differences in lifestyle risk ^42–45^ operate at different levels and cannot be separated with district-aggregated data alone. Before these rankings are used to direct resources, the further research on district-level indicators of deprivation would be helpful.

The steady decade-long rise in the ASPR of HTN and T2DM and their complications partly reflects the legacy of past increases in lifestyle risks prevalence. Because our estimates were standardized to a fixed WHO reference population, this upward trend cannot be explained by changes in age structure over time; it more plausibly reflects a combination of rising incidence ^46^, improved survival of prevalent cases ^47–49^, and intensified case detection through expanded primary-care screening over the study period ^50,51^. The SII results further support this conclusion. During the study period, the screening power was expanding, and the management rate was increasing with the policy implementation. Whatever the mix, the geographic pattern layered on top of this trend is spatially persistent, strengthening the case for treating high-burden districts as durable priorities for resource allocation rather than transient hotspots.

Our finding challenge optimism that the introduction of new treatments can address disparities. Because the deepest inequalities cluster in the low-prevalence, high-complication end-organ conditions (renal failure, heart failure, amputation) the districts falling behind are largely doing so at the stage of long-term management. The persistence of this heterogeneity across an entire decade, in conditions overwhelmingly driven by modifiable risk and care access, is hard to reconcile with the expectation that precision-medicine approaches will resolve the root causes of this burden. The top-ranked district still varied from one condition to the next. This partial, condition-specific clustering argues against a one-size-fits-all “target the worst districts” strategy and in favor of matching the intervention to the disease profile of each district. A district leading in HTN or T2DM prevalence in the general population calls for upstream, population-level prevention, whereas a district leading in complications within the cohorts calls for tighter secondary prevention and follow-up of patients already in care. The levers that plausibly move these gaps are population-level and behavioral: community programs for HTN centered on salt reduction and physical activity ^52,53^, and structured diabetes management such as the Risk Assessment and Management Program ^54^ for T2DM, deployed selectively to the districts where each condition peaks.

The identification of specific high-burden districts in the general population, and of conditions with robust geographic inequality, provides an empirical basis for district-specific public health planning. These findings argue for the equitable allocation of primary prevention strategies toward high-burden districts.

### Strengths

This study has several methodological strengths. The use of four complementary administrative databases with harmonized first-record dating and a carry-forward rule provides comprehensive, longitudinally consistent case ascertainment across the full 2014-2023 period and mitigates the under-counting that would arise from any single source. The inverse-attendance weighting addresses the attendance rate limitation of administrative data by rescaling observed cases to the resident population. The parallel use of direct standardization for high-prevalence conditions and Bayesian hierarchical modelling for rare conditions allows each disease to be analyzed with the method best suited to its data density. Applying the WHO standard across years ensures that the observed temporal increase is not confounded by changes in the reference population. Finally, the precision-weighted Cochran’s Q framework ensures that the heterogeneity measure operates directly on the age-standardized rates and appropriately down-weights imprecise estimates from small districts.

### Limitations

Several limitations warrant emphasis. The inverse-attendance weighting rests on the assumption that within each age-district-sex-year stratum disease prevalence among HA attendees equals that among non-attendees. Because attendees are, on average, sicker than non-attendees, this assumption may lead to over-estimation of true population prevalence; the direction and magnitude of any residual bias cannot be determined from these data alone. Second, district assignment relied on attendance-based encounters and a stability assumption embedded in the forward- and backward-fill imputation; patients whose true residence changed, or who attended outside their district of residence, may have been misassigned, and the 0.56% of patient-years left unassigned were excluded, potentially introducing selection. Third, disease ascertainment depended on the accuracy and completeness of clinical coding across four different datasets, and differential coding practices across districts could manifest as apparent geographic heterogeneity. Fourth, as an ecological and cross-sectional-by-year analysis, the study cannot support causal inference about the determinants of the observed district inequalities. Finally, fitting a slope across 18 districts yields limited statistical power and the small number of areas also means the PCA loadings, while substantively coherent, are estimated with limited precision.

## Conclusion

Using territory-wide linked administrative records and a combination of direct and Bayesian standardization methods, we found significant and statistically robust geographic disparities in the prevalence of HTN, T2DM, and their complications across Hong Kong’s eighteen districts, against a backdrop of rising burden from 2014 to 2023. The concentration of high prevalence in districts such as Kowloon City and Yuen Long underscores the need for district-specific public health strategies, equitable allocation of primary-care resources, and targeted chronic-disease prevention and management programs in high-burden areas. District-level prevalence of T2DM and diabetic retinopathy is higher in socioeconomically disadvantaged districts of Hong Kong, and these inequalities widened between 2014 and 2023. The gradients are consistent across four measures of socioeconomic position. Notwithstanding the limited precision inherent in an 18-area design, these findings support the geographic targeting of cardiometabolic prevention and screening toward lower socioeconomic-position districts and illustrate the value of testing socioeconomic-inequality findings against multiple operationalization of socioeconomic position.

## Author Contributions

Conceptualization: Jingjing Zhou, David Bishai.

Data curation: Jingjing Zhou, Zhenyuan Liu, Qiaohui Wu.

Formal analysis: Jingjing Zhou.

Funding acquisition: David Bishai.

Methodology: Jingjing Zhou, Yuchen Cai, David Bishai.

Project administration: Jingjing Zhou, David Bishai.

Resources: David Bishai.

Software: Jingjing Zhou.

Supervision: David Bishai.

Validation: Jingjing Zhou, Zhenyuan Liu, Qiaohui Wu.

Visualization: Jingjing Zhou.

Writing – original draft: Jingjing Zhou, David Bishai.

Writing – review & editing: Jingjing Zhou, Zhenyuan Liu, Yuchen Cai, Qiaohui Wu, David Bishai.

## Ethics Approval and Consent to Participate

This study was approved by the Institutional Review Board of the University of Hong Kong/ Hospital Authority Hong Kong West Cluster (Approval Number: UW 24-109). All procedures followed data privacy protection and ethical guidelines.

## Data Availability

The data underlying the results presented in the study are available from the Hong Kong Hospital Authority Data.

## Acknowledgments

The authors thank the HA of Hong Kong for kindly providing the data.

## Funding

This work was supported by the Health and Medical Research Fund (HMRF). The funder had no role in the study design, data collection and analysis, decision to publish, or preparation of the manuscript.

## Conflict of Interest

The authors declare no conflict of interest.

## Supplementary Dashboard

Age-Standardized Comorbidity Prevalence across Total, Diabetic, and Hypertensive Populations

**Supplementary Table 1.** Diseases codes used in the model by Diabetes Mellitus Complications Surveillance (DMCS), International Classification of Disease (ICD-9) and International Classification of Primary Care (ICPC-2) coding

| Outcomes | DMCS | ICD-9 | ICPC-2 |
| --- | --- | --- | --- |
| Coronary heart disease | echd_flag: Y<br>chd_flag:: Y | 410-414 | K74-K76 |
| Heart failure |  | 402.01, 402.11, 402.91,<br>404.01, 404.03, 404.11,<br>404.13, 404.91, 404.93,<br>428.0, 428.1, 428.9 | K77 |
| Cerebrovascular disease | stroke_hx_flag: Y<br>cva_unk_flag: Y<br>stroke_flag: Y | 430-438 | K90-K91 |
| Atrial fibrillation |  | 427.31-427.32 | K78 |
| Neuropathy |  | 250.6, 250.62, 337.0,<br>337.1, 355, 356, 357 | N94 |
| Cataract | pres_cataract_r_flag: Y<br>pres_cataract_l_flag: Y<br>cataract_ext_r_flag: Y<br>cataract_extr_l_flag: Y | 366 | F92 |
| Retinopathy | retina_sum_cd: Y<br>risk_ht_retin_flag: Y<br>risk_diab_retin_cd: Y | 362 | F83 |
| Peripheral vascular diseases |  | 250.7x, 440, 443.1-443.2, 443.8-443.9,<br>785.4, 997.2<br>Procedure code:<br>39.29, 39.9 | K92 |
| Skin ulcer or amputation of<br>lower limb | hist_ulcer_amp_flag: Y | 707, 997.6x<br>Procedure code 84.11 | S97 |
| Renal failure |  | 403.01, 403.11, 403.91,<br>404.02, 404.03, 404.12,<br>404.13, 404.92, 404.93,<br>585-586<br>Procedure code 39.95 |  |
| Hyperlipidemia | dyslipidaemia_flag: Y | 272.x | T93 |
| Hypertension | htn_flag: Y<br>ht_dx_date_y | 401 | K85, K86, K87 |
| Type 2 Diabetes Mellitus | dm_type: Type 2<br>diabetes_cd: D/I/Y<br>dm_dx_date_y | 250.00, 250.02, 250.10,<br>250.12, 250.20, 250.22,<br>250.30, 250.32, 250.40, | T90 |
|  |  | 250.42, 250.50, 250.52,<br>250.60, 250.62, 250.70,<br>250.72, 250.80, 250.82,<br>250.90, 250.92 |  |

DMCS: Diabetes Mellitus Complications Surveillance; ICD-9: International Classification of Diseases Clinical Modification; ICPC-2: International Classification of Primary Care, second edition; N94 and S97 have no records.

## Notes

### Competing Interest Statement

The authors have declared no competing interest.

